# The impact of pet care challenges on medical decision making and healthcare utilization among hospitalized pet owners: a prospective study

**DOI:** 10.64898/2026.01.16.26344223

**Authors:** Carri S. Polick, Jennifer W. Applebaum, Joshua Errickson, Ronald D. Chervin, Megan A. Iida, Nikki Chiang, David Johnson, Doug Plant, Tiffany J. Braley

**Affiliations:** School of Nursing, Duke University, Durham, NC, USA; Department of Environmental and Global Health, University of Florida, Gainesville, FL, USA; Consulting for Statistics, Computing and Analytics Research (CSCAR), University of Michigan, Ann Arbor, MI, USA; Department of Neurology, University of Michigan - Michigan Medicine, Ann Arbor, MI, USA; Department of Neurology, University of California San Francisco, San Francisco, CA, USA; Department of Pathology, University of Michigan - Michigan Medicine, Ann Arbor, MI, USA; Department of Physical Medicine and Rehabilitation - Michigan Medicine, Ann Arbor, MI, USA; Michigan Humane, Detroit, MI, USA

## Abstract

**Background:** Little is known about the scope of challenges faced by hospitalized pet owners and their impact on medical decision-making and utilization of health services.

**Objectives:** To assess challenges associated with pet care (PC) among hospitalized pet owners and characterize associations between reported challenges, patient characteristics, and adherence to treatment.

**Methods:** Pet owners in inpatient or ED settings were surveyed about PC challenges experienced during current and prior hospitalizations, and social support for PC. Electronic medical records were followed prospectively. Bivariate tests and logistic/multiple linear regression models were used to examine associations between survey responses, diagnoses, and healthcare utilization.

**Results:** Among 352 respondents (71 ED, 281 inpatient), younger age (p<0.001), less social support (p<0.01), and higher comorbidity (p=0.03) were associated with more PC challenges. Twenty-five ED patients (35%) reported challenges, of whom 46% endorsed likely deferral of hospital admission if recommended (p<0.0001). N=67 inpatients (24%) endorsed PC challenges during their current hospitalization. Prior challenges predicted current challenges (r=0.34, p<0.001). Increased social support reduced the odds of PC challenges (OR:0.80, p<0.01).

**Conclusions:** Pet care challenges are common among pet owners, who report substantial impact on medical decision-making and healthcare utilization. Those with reduced social support and prior challenges are particularly vulnerable.

## INTRODUCTION

Approximately 60% of US households include pets^1^. Although pet ownership has been shown to positively impact human health and wellbeing^2,3^, pet care responsibilities have the potential to hinder healthcare utilization for owners who need Emergency Department (ED) or inpatient care. Lack of social support to assist with pet care could also contribute to deviations from recommended treatment plans. Presently, hospitals lack formalized plans or programs to assist with pet care, which could improve patients’ ability to accept inpatient care, particularly for extended treatments such as inpatient rehabilitation. Partnerships with foundations and animal welfare groups, informed by data regarding the scope of impact, greatest needs, and patient preferences, could support this need.

Evidence suggests that pets frequently factor into healthcare decision-making^4,5^. In a prior survey study of patient representatives and their families, 63% reported that they or one of their loved ones had prior challenging experiences securing pet care during hospitalizations, with most reporting that these challenges were linked to negative consequences for recovery or medical decision-making (MDM)^6^. This foundational retrospective research highlighted pet care as a new facet of MDM. Yet, research that captures pet owner judgement during real-life medical situations, and potential consequences of altered MDM, is lacking.

Hospital courses and public health could potentially be optimized by a better understanding of how pet caregiver status affects adherence to medical recommendations, recovery, and utilization across medical settings. However, additional information is first needed to understand which patients are most vulnerable to pet care challenges and associated health consequences. Prior work suggests that younger and non-binary gender patients may have increased pet care challenges; however evidence to date regarding these populations is limited^6^.

Although vulnerable clinical populations are also under-explored, prior research suggests that pet owners with HIV, especially those with poor social support, may be vulnerable to treatment plan interruptions secondary to pet care responsibilities^7^. Acute conditions that require emergent hospitalization (e.g., stroke) could also be associated with difficulty securing pet care, but these experiences remain unstudied.

The overarching goals of this study were to determine the scope of pet care challenges experienced by ED and hospitalized patients at a tertiary care center, identify subgroups at higher risk for challenges, and characterize associations between pet care challenges and healthcare utilization (e.g., readmissions). We hypothesized that emergent conditions, lower social support, and non-binary gender would be associated with increased pet care challenges, and difficulties with MDM regarding healthcare utilization.

## METHODS

### Respondents/data collection procedures

The University of Michigan Institutional Review Board (IRB-MED) approved all study activities prior to study commencement. All participants and/or their legal guardian(s) provided their informed consent to participate in the study. Study activities were performed in accordance with relevant named guidelines and regulations.

Participants included adults either 1) seeking ED care or 2) admitted to an inpatient unit at Michigan Medicine. Participants had to identify as a caregiver for at least one pet. Eligible individuals were invited to consent to an online survey and medical record review.

Participants were approached by the study team or notified through flyers posted throughout the hospital. Using QR codes to a secure online platform^8^, surveys were completed by patients with their smart device, or with study team tablets. Since COVID-19-imposed restrictions curtailed in-person research, the vast majority of surveys were collected after restrictions were lifted (2022-2023). Efforts were made to over-sample several participant subgroups to support racial and medical subgroup analyses (e.g., those admitted with primary neurological diagnoses).

Electronic medical records (EMR) were reviewed from admission to 90-days post-discharge. Zip codes were used to retrieve 2023 census track median household income. EMR data included visit diagnosis, length of stay, surgical procedure urgency, discharge disposition, presence of ED visit within 30-days, and inpatient readmission within 30 and 90-days. Potential indicators of challenges such as Social Work consultations were also assessed. Encounter summaries differed by reporting Charlson^9^ or Elixhauser^10^ Comorbidity Index scores. These were compared to the number of active problems, which were strongly correlated and covaried in adjusted models.

### Survey content

Two location-specific surveys were developed. Overlapping items included pet type/number, social support available to assist with pet care, experiences with prior pet care challenges in medical settings and the impact on care. All participants were queried regarding perceived value of screening and referral to low/no cost resources (e.g., boarding, fostering), and whether visiting with their pet would be helpful to their recovery. The ED version included items regarding opportunity for pet care preparation before seeking ED care, and potential availability of online caregiving resources. For example, participants coming from a clinic or scene (e.g., car accident) may lack such opportunities compared to participants who left from home. As ED participants may not have known whether they were going to be admitted, a hypothetical question assessed their anticipated plan if hospitalization was recommended.

### Pet care challenge survey items

Both ED and inpatient surveys asked about the presence of challenges in finding someone to assist with pet care during a prior hospital admission (if applicable), and whether pet care challenges ever influenced a decision to receive any type of medical care, truncate a hospital stay, forego a procedure, or accept a hospital admission. The inpatient sample was also asked about challenges during the *current* admission. Responses for these six variables were collapsed into a composite that reflected the presence of any type of challenge at any time. This composite was used for analytic models that involved the entire sample. To better evaluate challenges relating to current and past admissions, additional models were tailored using a binary variable on *challenges during current admission* within the inpatient subsample. Similarly, we restricted the variable regarding presence of a *challenge during prior admission* by removing participants who reported no prior admissions.

### Social support for pet care

Complexities in habitation and social situations may result in varying levels of support available for pets. To capture these nuances, we composed a Social Score count consisting of six variables to represent the latent concept of a social support network for pet care. Social scores ranged from 0-6. Specifically, participants received one-count for each affirmative response: 1) Being married, 2) Living with spouse/domestic partner, 3) Spouse/partner assistance with pet care, 4) Family/friend assistance with pet care, 5) Living with other family, and 6) Living with roommates.

### Emergent/Complex Diagnoses

Two reviewers coded primary discharge diagnoses independently to capture emergent and/or medically-complex situations that were more likely to result in severe morbidity or death in the absence of immediate medical attention. Alignment was moderately high (Cohen’s Kappa 0.78), and discrepancies resolved with discussion.

### Qualitative answers

If a participant endorsed a challenge on a particular item, they were given the opportunity to elaborate with free-text qualitative responses. Responses were organized into themes and sub-themes using a realist thematic analysis approach, typically used to report experiences and the reality of participants^11^. Responses were themed by two reviewers with high alignment and any differences easily settled with discussion.

### Statistical methods

Pearson correlations, Fisher’s exact tests, and Chi-Square tests were used to examine bivariate associations between participant characteristics, experience with pet care challenges, and hospital outcomes that informed regression models.

Logistic regression models were constructed to first examine associations between demographic/clinical characteristics (primary independent variables of interest) and three binary (yes/no) pet care challenge outcomes: 1) composite of prior and/or current challenges, 2) challenges during a *prior* admission, and 3) challenge during the *current* admission.

Separate logistic and multiple linear regression models were then constructed to examine associations of *prior* and *current* pet care challenges (as separate independent variables) and four hospital outcomes chronologically including the length of current hospital stay, presence of 30-day ED visit, and presence inpatient readmission at 30- and 90-days post-discharge.

## RESULTS

### Baseline characteristics

After removing friends/family who responded to the flyer, 910 patients were screened for eligibility (**Figure 1**). Of those, 57.4% (n=522) were pet owners. Of pet owners, 12.5% (n=65) were not interested, and 17.8% (n=93) did not complete screening/consent. Overall, 352 patients had an encounter for chart review and were included in initial analyses. Most (n=345) could be tracked for the full 90-days and were included in final analyses (e.g., return visits).

**Figure 1:**
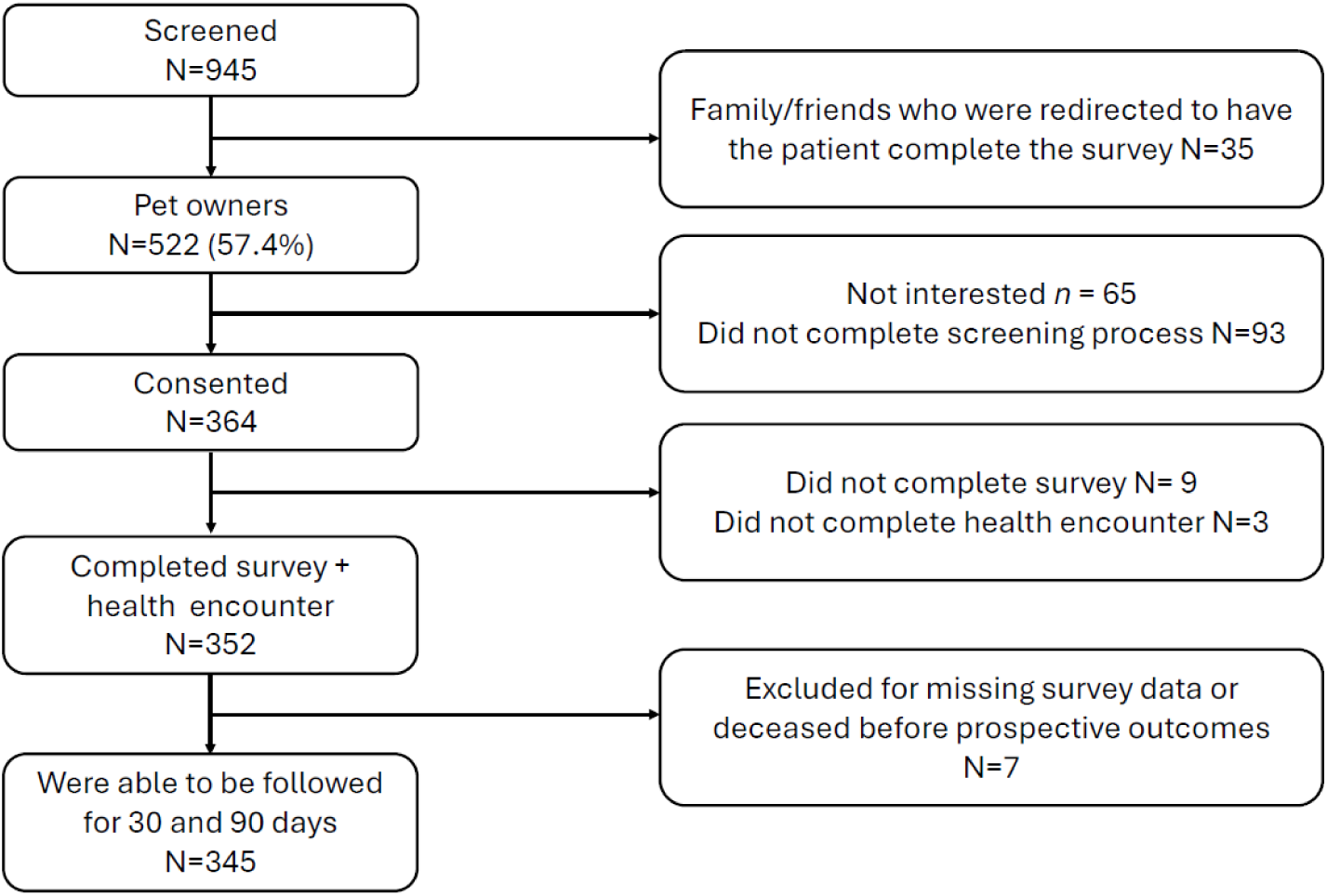
Participant Flow

Participants were mostly women (54%), White (87%), hospitalized (80%), had at least one dog (75%), and median household income was $73,000 (**Table 1**). Most participants had a spouse/significant other (70%) and family, friends, or neighbors (88%) who could be called upon to provide pet care. ED and inpatient subsample characteristics did not significantly differ except regarding age (49.9 vs 56.9, respectively, p=0.001), mean comorbidity count (9.2 vs 11.1, p<0.05), and mean length of stay (79.7 vs 292.9 hours, p=0.019). Relative to White participants, Black participants had fewer household pets (r= -0.11, p=0.03) (**Supplementary Table 1**).

**Table 1.** Participant characteristics, challenges, and program preferences by hospital setting.

|  |  |  |  |  |
| --- | --- | --- | --- | --- |
|  | **Total** | **Inpatient Units** | **Emergency Department** | **Test** |
|  | (N=352) | (N=281) | (N=71) | p-value |
| Age (n=351) | 55.5 (16.5) | 56.9 (16.5) | 49.9 (15.3) | 0.001* |
| Gender (n=352) |  |  |  |  |
| Women | 190 (54.0%) | 143 (50.9%) | 47 (66.2%) | 0.063 |
| Men | 152 (43.2%) | 130 (46.3%) | 22 (31%) |  |
| Non-binary | 10 (2.8%) | 8 (2.8%) | 2 (2.8%) |  |
| Race (n=352) |  |  |  |  |
| White | 305 (86.6%) | 242 (86.1%) | 63 (88.7%) | 0.775 |
| Black | 26 (7.4%) | 21 (7.5%) | 5 (7%) |  |
| Other | 21 (6.0%) | 18 (6.4%) | 3 (4.2%) |  |
| Median household Salary (in \$1,000 increments) (n=352) | 73 (23.5) | 72.7 (24.1) | 74.3 (21) | 0.608 |
| # of active problems (n=352) | 10.7 (7.2) | 11.1 (7) | 9.2 (7.9) | 0.048* |
| Has dog(s) (n=352) | 264 (75%) | 214 (76.2%) | 50 (70.4%) | 0.319 |
| Has cats(s) (n=352) | 162 (46%) | 127 (45.2%) | 35 (49.3%) | 0.536 |
| Has other pets(s) (n=352) | 57 (16.2%) | 46 (16.4%) | 11 (15.5%) | 0.858 |
| Has significant other/spouse who helps with pet(s) (n=299) | 208 (69.6%) | 163 (70.6%) | 45 (66.2%) | 0.490 |
| Has nearby family/friends/others who could help (n=298) | 263 (88.3%) | 207 (90%) | 56 (82.4%) | 0.085 |
| **Challenges securing pet care** |  |  |  |  |
| Had challenge finding someone to care for pets during **current** inpatient hospitalization (n=276) | - | 67 (24.3%) | - | - |
| Had challenge during **prior** hospitalizations (n=266)<sup>a</sup> | 60 (22.6%) | 44 (20.7%) | 16 (30.2%) | 0.137 |
| Decision to receive any type of recommended medical care was ever impacted by a pet care challenge (n=324) | 48 (14.8%) | 35 (13.3%) | 13 (21.3%) | 0.113 |
| Decision to shorten hospital stay was ever impacted by a pet care challenge (n=309) | 23 (7.4%) | 17 (6.6%) | 6 (11.3%) | 0.237 |
| Decision to forego procedure was ever impacted by a pet care challenge (n=328) | 22 (6.7%) | 15 (5.6%) | 7 (11.5%) | 0.099 |
| Decision to accept admission was ever impacted by a pet care challenge (n=329) | 26 (7.9%) | 19 (6.9%) | 7 (12.7%) | 0.146 |
| Binary composite of at least one type of challenge above (during current and/or prior hospitalization) (n=345) | 132 (38%) | 107 (38.2%) | 25 (37.3%) | 0.891 |
| **ED specific items** |  |  |  |  |
| Made a plan for pet when deciding to seek ED care (n=68) | - | - | 43 (63.2%) | - |
| Where were you when you left for the ED? (n=68) |  |  |  | - |
| Home | - | N/A | 52 (76.5%) | - |
| Hospital Clinic | - | - | 5 (7.4%) | - |
| Work | - | - | 5 (7.4%) | - |
| From a scene | - | - | 3 (4.4%) | - |
| Other | - | - | 3 (4.4%) | - |
| How likely do you think it is that you will get admitted to the hospital? (n=68) |  |  |  |  |
| I know I will get admitted | - | - | 19 (27.9%) | - |
| I think I might get admitted | - | - | 14 (20.6%) | - |
| I don't know | - | - | 8 (11.8%) | - |
| I think I will be discharged after seeing a doctor | - | - | 27 (39.7%) | - |
| If you were told that you needed to be admitted to the hospital, would your pet(s) influence your decision to stay? (n=68) |  |  |  |  |
| Yes, I would consider going home against medical advice if I couldn't secure pet care | - | - | 11 (16.2%) | - |
| I don't know | - | - | 4 (5.9%) | - |
| No, I will follow the doctor's admission advice even if I can't find a caregiver for my pet(s) | - | - | 3 (4.4%) | - |
| No, I have a plan for pet care already | - | - | 50 (73.5%) | - |
| Had ever used services like Rover or Care.com (n=70) | - | - | 9 (13.2%) | - |
| **Programmatic preferences** |  |  |  |  |
| Think there should be a low/no cost program to assist hospitalized patients with pet care (n=298) | 293 (98.3%) | 225 (97.8%) | 68 (100%) | 0.220 |
| Would consider using pet boarding program (n=298) | 248 (83.2%) | 188 (81.7%) | 60 (88.2%) | 0.208 |
| Would consider using pet foster program (n=298) | 240 (80.5%) | 182 (79.1%) | 58 (85.3%) | 0.259 |
| Pet visit would be helpful to their recovery (n=298) | 226 (75.8%) | 178 (77.4%) | 48 (70.6%) | 0.250 |
| **Hospital utilization** |  |  |  |  |
| Social work referral placed for any reason (n=351) | 151 (42.9%) | 137 (48.8%) | 14 (19.7%) | - |
| Was inpatient rehabilitation recommended (n=351) | 27 (7.7%) | 26 (9.3%) | 1 (1.4%) | - |
| Did patient pursue inpatient rehabilitation (n=351) | 23 (6.6%) | 22 (7.8%) | 1 (1.4%) | - |
| Left AMA (n=351) | 5 (1.4%) | 4 (1.4%) | 1 (1.4%) | - |
| Length of stay (hours) M(SD) (n=317) | 268.7 (516.1) | 292.9 (542.6) | 79.7 (89.1) | 0.019* |
| Had ER revisit within 30 days (n=351) | 80 (22.8%) | 60 (21.4%) | 20 (28.2%) | 0.227 |
| Was readmitted to hospital within 30 days (n=351) | 75 (21.4%) | 62 (22.1%) | 13 (18.3%) | 0.482 |
| Was readmitted to hospital within 90 days (n=351) | 113 (32.3%) | 96 (34.3%) | 17 (23.9%) | 0.096 |
| *p<0.05 |  |  |  |  |
| - Denotes that testing between ED and inpatient sample not available or applicable |  |  |  |  |

<sup>a</sup>Excludes participants who are admitted for the first time and do not have a prior hospitalization

### Pet care challenges and MDM

Over a third (n=132, 38%) of participants endorsed challenges in securing pet care that impacted either current or prior MDM. Twenty-three percent (n=60) of the total sample who experienced at least one prior hospitalization reported challenges during these prior admissions, while 24% (n=67) of the inpatient sample had challenges during their *current* admission. Pet care challenges had reported impact on decisions to receive any type of recommended treatment (n=48, 15%), shorten a hospital stay (n=23, 7%), forego a procedure (n=22, 7%), and accept admission (n=26, 8%). Optional qualitative responses highlighted personal experiences with difficult decisions and adverse outcomes that included jeopardized human/pet wellbeing and pet re-homing (**Table 2**).

**Table 2.** Participant descriptions of pet care needs impact on medical decisions and experiences (n= 76)

|  |  |
| --- | --- |
| **Theme: Deferral, delay, or truncation of care** |  |
| Example 1 | “I left instead of getting admitted” |
| Example 2 | “Cancelled scheduled surgery due to inability to get dog care” |
| **Sub-theme: Deferring inpatient rehabilitation (n=6)** | “Always try to get out as soon as I am stable and always refuse rehab due to pets. I am their mother and they are my responsibility.” |
| **Sub-theme: Delaying medical treatment (n=26)** | “I needed to come to the hospital when my partner and local family were out of town, but chose to delay treatment because I didn't have help to care for my dog.” |
| **Sub-theme: Incomplete medical Care (n=27)** | “I was admitted and left before [a feeding tube procedure] to care for my cat after two days when I was told I'd need to stay another 3 days at least.” |
| **Theme: Negative outcomes for patient and/or pet (n=12)** |  |
| Example 1 | Dog died of neglect under temporary caregiver while patient was admitted |
| Example 2 | Care team recommended staying but patient left and reported regretting it since they needed to return to hospital for additional medical care |
| **Theme: Other patient views/experiences** |  |
| Example 1 | “My dog is a very important part of my mental health and I am trying to get her registered as an emotional support dog so she can be in the hospital with me.” |
| Example 2 | “It a huge burden to put on somebody else. Nobody takes care of your pets like you do.” |
| **Sub-theme: Near misses (n=4)** | “Considering leaving earlier during this stay, then cousin stopped by” |

### ED subsample

Most ED participants left from home (n=52, 77%) and had already planned for pet care (n=43, 63%). Among the 68 participants who shared their anticipated response to a hospital admission recommendation, 16% (n=11) reported they would consider going home against medical advice (AMA) if they couldn’t secure pet care. Participants who presented with emergent medical conditions were more likely to experience challenges. Participants who were able to make plans before seeking ED care mostly reported that pet care would not impact their decision to stay for admission.

Post hoc Fisher’s exact testing revealed that anticipated response to an admission recommendation significantly differed by previous experience with a pet care challenge. Two thirds of participants who considered declining admission if they lacked pet care support had experienced at least one previous challenge (n=12, 75%). In contrast, 75% of those who reported that pets would not impact their decision to accept admission had not previously experienced a pet care challenge (n=39, p=0.001). **Supplementary materials** highlight correlations between current inpatient challenges and challenges during prior hospitalizations (r=0.34, p<0.001), and between challenges and MDM about accepting (r=0.17, p<0.01) and shortening (r=0.24, p<0.001) admissions.

### Program Building

The vast majority (n=293, 98%) thought that a low/no cost pet care program should be offered to hospitalized patients without sufficient social or financial resources (**Table 1**).

### Associations between patient characteristics and challenges

In three adjusted models that examined associations between patient characteristics and pet care challenges, younger age and increased number of comorbidities were consistently associated with prior challenges and combined challenges (any prior/current challenge) that impacted MDM. This trend was not significant regarding only current challenges in the inpatient subsample (**Table 3**). Each additional year aged was associated with 3% lower odds of experiencing prior or combined challenges (both model ORs 0.97, p<0.01), such that a 20-year-old would have 90% higher odds of pet care challenges compared to a 50-year-old. Each additional active comorbidity was associated with 4% higher odds of having any type of challenge (OR 1.04, p=0.03) and 9% higher odds of challenges during prior hospitalizations (OR 1.09, p<0.001).

**Table 3:** Predictors of prior or current pet care challenges which impacted medical decision making.

|  |  |  |  |
| --- | --- | --- | --- |
|  | **Challenges ever**<br>**(prior and/or current)**<br>**(n=346)** | **Challenges during**<br>**prior admissions**<br>**(n=251)** | **Challenges during current**<br>**admission**<br>**(inpatient sample only)**<br>**n=275** |
| # of active medical problems | 1.04 (0.02),<br>**0.03\*** | 1.09 (0.02),<br>**<0.001\*\*** | 1.01 (0.02),<br>0.68 |
| Age | 0.97 (0.01),<br>**<0.001\*\*** | 0.97 (0.01)<br>**<0.01\*\*** | 1.00 (0.01),<br>0.62 |
| Gender (ref. male) |  |  |  |
| Female | 0.84 (0.21),<br>0.49 | 1.71 (0.63),<br>0.15 | 0.77 (0.23),<br>0.37 |
| Non-binary | 3.30 (2.78),<br>0.16 | 6.56 (5.31),<br>**0.02\*** | 0.90 (0.81),<br>0.91 |
| Race (ref. White) |  |  |  |
| Black | 0.32 (0.17),<br>**0.03\*** | N/A<sup>b</sup> | 0.28 (0.22),<br>0.10 |
| All others combined | 0.70 (0.38),<br>0.51 | 0.95 (0.71),<br>0.94 | 0.64 (0.43),<br>0.51 |
| Social Score (social support to help with pet care) | 0.80 (0.06),<br>**<0.01\*\*** | 0.82 (0.09),<br>0.08 | 0.96 (0.9),<br>0.71 |
| Median Income (zip code) | 0.97 (0.05),<br>0.52 | 0.93 (0.07),<br>0.32 | 0.98 (0.06),<br>0.73 |
| Total pets (ref. 1) |  |  |  |
| 2 | 0.73 (0.21),<br>0.29 | 1.39 (0.58),<br>0.43 | 0.91 (0.32),<br>0.79 |
| 3 or more | 0.99 (0.28),<br>0.97 | 1.77 (0.74),<br>0.17 | 1.12 (0.39),<br>0.75 |
| Emergent/severe primary diagnosis (ref. No) |  |  |  |
| Yes | N/A<sup>a</sup> | N/A<sup>a</sup> | 1.54 (0.46),<br>0.15 |
| Intercept | 5.41 (3.68),<br>**0.01\*** | 0.90 (0.83),<br>0.91 | 0.50 (0.41),<br>0.39 |
| Pseudo R-squared | 0.08 | 0.17 | 0.03 |
| All estimates listed as OR (SE), p-value, ** p<0.01, * p<0.05 |  |  |  |
| <sup>a</sup>Not applicable for past admissions |  |  |  |
| <sup>b</sup>Not available. All Black subgroup participants who answered this question were n/a for not having a prior admission (n=9), or denied challenges (n=14), thus estimates could not be calculated. |  |  |  |

In bivariate analyses, social support for pet care significantly correlated with fewer challenges (**Supplementary Table 1**), except current inpatient challenges which remained negatively correlated, but non-significant. In adjusted models, each additional level of social support was associated with 20% lower odds of having a prior and/or current challenge (OR 0.8, p=0.01, **Table 3**). Black participants had 68% lower odds of ever experiencing pet care challenges compared to White participants (OR 0.32 p=0.03). Number of pets did not significantly predict pet care challenges.

A trend toward significance was seen among pet care challenges and emergent diagnoses in Pearson correlation tests (r=0.10, p=0.09, **Supplementary Table 1**).

Regarding challenges during prior hospitalizations, non-binary gender participants (n=10) had nearly seven times higher odds of experiencing pet care challenges compared to men (OR 6.56, p=0.02). However, smaller numbers of participants were noted across some race and gender categories, thus caution should be applied to interpretation.

### Service Utilization and Disposition

Forty-three percent of participants (n=151) had a social work consultation, which was more common for the inpatient setting (n=137; **Table 1**). Those who endorsed that pet care challenges had impacted MDM (regarding any type of treatment) were more likely to have a social work consult while admitted (r=0.16, p=0.004) (**Supplementary Table 1**). Having a social work consult was also associated with higher 30-day readmission rates (r=0.11, p=0.036).

Twenty-three of 27 patients recommended for inpatient rehabilitation accepted it (**Table 1**). Three of the four who deferred had experienced pet care challenges (current/past).

Rehabilitation acceptance showed a trend toward association with reports of current pet care challenges (r=0.11, p=0.058, **Supplementary Table 1**). In subgroup analyses, current pet care challenges positively correlated with inpatient rehabilitation (r=0.14, p=0.03) among non-neurology patients (**Supplementary Table 2**), but these correlations were not significant in the neurology subgroup..

In bivariate analyses non-binary gender participants, in comparison to others, had more pets (r=0.15, p<0.01), less social support (r= -0.14, p=0.01), more pet care challenges that impacted decisions to receive any treatment (r=0.19, p=0.001), and higher likelihood of leaving AMA (r=0.17, p<0.01). Although few patients overall had a documented AMA discharge (n=5), additional patients had similar, less stigmatized, chart language (e.g., “very eager to leave”). Adjusted analyses revealed that patients with current pet care challenges have 40% longer admissions than patients without challenges (p=0.01; **Table 4**).

**Table 4.** Prospective healthcare utilization outcomes.

| **Table 4. Prospective healthcare utilization outcomes** |  |  |  |  |  |  |  |
| --- | --- | --- | --- | --- | --- | --- | --- |
| Independent variable of interest and covariates | Dependent variables, regressed on any challenges, prior or current (full sample $n=345$) | | | Dependent variables, regressed on challenges during current admission (inpatient sample only, $n=274$ for 30- and 90-day outcomes) | | | |
| | 30 day ER visit | 30 day readmit | 90 day readmit | <sup>a</sup>Length of hospital stay ($n=275$) | 30 day ER visit | 30 day readmit | 90 day readmit |
| **Challenge Yes** (ref. No) | 1.06 (0.31)<br>0.85 | 1.06 (0.31),<br>0.84 | 0.87 (0.22),<br>0.58 | 39.9%<br>0.34 (0.13),<br>**0.01\*** | 0.75 (0.28),<br>0.46 | 1.36 (0.47),<br>0.37 | 1.31 (0.40),<br>0.38 |
| **# of active problems** | 1.06 (0.02)<br>**0.001\*\*** | 1.06 (0.02),<br>**<0.01\*\*** | 1.05 (0.02),<br>**<0.01\*\*** | 1.4%<br>0.01 (0.01),<br>0.09 | 1.06 (0.02),<br>**<0.01\*\*** | 1.05 (0.02),<br>**0.03\*** | 1.05 (0.02),<br>**<0.01\*\*** |
| **Age** | 1.00 (0.01),<br>0.96 | 0.99 (0.01),<br>0.54 | 1.00 (0.01),<br>0.82 | 0.2%<br>0.002, (0.00),<br>0.59) | 1.00 (0.01),<br>0.72 | 0.99 (0.01),<br>0.40 | 1.00 (0.01),<br>0.76 |
| **Gender** (ref. male) |  |  |  |  |  |  |  |
| Female | 1.20 (0.34),<br>0.51 | 1.31 (0.37),<br>0.34 | 1.10 (0.27),<br>0.71 | -17.2%<br>-0.19 (0.12),<br>0.11 | 1.35 (0.43),<br>0.35 | 1.59 (0.50),<br>0.14 | 1.21 (0.34),<br>0.48 |
| Non-binary | 1.47 (1.14),<br>0.62 | 1.24 (1.07),<br>0.81 | 2.30 (1.66),<br>0.25 | -38.1%<br>-0.48 (0.35), | 3.61 (3.08),<br>0.13 | 0.74 (0.85),<br>0.79 | 2.10 (1.17),<br>0.36 |
|  |  |  |  | 0.18 |  |  |  |
| **Race (ref. White)** |  |  |  |  |  |  |  |
| Black | 1.98 (0.95),<br>0.16 | 1.53 (0.77),<br>0.41 | 2.25 (1.02),<br>0.07 | -10.4%<br>-0.11 (0.23),<br>0.63 | **3.34** (1.74),<br>**0.02\*** | 2.52 (1.36),<br>0.09 | 3.75 (1.93),<br>**0.01\*** |
| All others combined | 0.79 (0.53),<br>0.73 | 0.20 (0.21),<br>0.13 | 0.39 (0.26),<br>0.15 | -0.8%<br>-0.01, (0.24),<br>0.97 | 0.51 (0.41),<br>0.40 | 0.21 (0.22),<br>0.14 | 0.44 (0.30),<br>0.22 |
| **Social Score** | 0.96 (0.09),<br>0.69 | 1.06 (0.10),<br>0.55 | 1.14 (0.09),<br>0.11 | 6.8%<br>0.07 (0.04),<br>0.08 | 1.06 (0.11),<br>0.56 | 1.08 (0.11),<br>0.45 | 1.16 (0.11),<br>0.10 |
| **Median Salary (zip code)** | 0.90 (0.06),<br>0.10 | 1.07 (0.06),<br>0.23 | 1.09 (0.06),<br>0.10 | -0.5%<br>-0.01 (0.02),<br>0.84 | 0.93 (0.06),<br>0.27 | 1.12 (0.07),<br>0.06 | 1.16 (0.07),<br>**0.01\*** |
| **Total pets (ref. 1)** |  |  |  |  |  |  |  |
| 2 | 1.43 (0.46),<br>0.26 | 0.78 (0.26)<br>0.45 | 0.82 (0.24),<br>0.50 | 8.5%<br>0.08 (0.14),<br>0.55 | 1.27 (0.45),<br>0.51 | 0.82 (0.30),<br>0.57 | 0.78 (0.25),<br>0.45 |
| 3 or more | 1.06 (0.36),<br>0.86 | 0.63 (0.21),<br>0.17 | 0.67 (0.20),<br>0.18 | -6.8%<br>-0.07 (0.14),<br>0.62 | 0.70 (0.28),<br>0.38 | 0.78 (0.29),<br>0.50 | 0.72 (0.24),<br>0.31 |

## DISCUSSION

To our knowledge, this study is the first to characterize the scope of pet care challenges experienced by hospitalized patients during their admissions or ED visits, revealing underexplored relationships between pet care challenges and healthcare utilization. Our findings suggest a high unmet need for pet care assistance, with potential to influence MDM among hospitalized pet owners. Our findings also illuminate several patient subgroups who may have an elevated risk of experiencing pet care challenges that affect their ability to accept recommended treatment.

A noteworthy proportion of participants experienced challenges securing pet care across both previous hospitalizations (22.6%) and current admissions (24.3%), and those who reported prior challenges were at higher risk of experiencing current challenges. Further, pet care challenges were associated with several aspects of MDM planning, including treatment adherence, delayed/deferred procedures, deferred admission, and AMA departures. Emergent situations, where patients may not have the choice or ability to leave, may risk adverse pet wellbeing outcomes such as those highlighted in our qualitative responses. If these findings can be generalized to other populations, people who have strong, health-promoting bonds with their pets could face difficult choices between their pets’ welfare and their own health, particularly when resources are constrained^12^. The human-animal bond has socio-emotional, health, economic, housing, and resource access implications, and can arguably be viewed as a social determinant of health for pet owners^6,13–16^. Our findings provide further support, showing that pet care responsibilities influence MDM and healthcare access. Many hospital systems have recently adopted routine social determinants of health screening^17,18^. This study aligns with broader goals to identify health-related social needs and provide support, including referral to community resources (e.g. community-based pet foster programs).

Pet care challenges may increase hospitalization-related stress and reduce treatment plan adherence. We found that nearly half of ED pet owners who reported pet care challenges indicated that they would consider leaving AMA if they could not secure pet care for an admission, while only 5% of ED pet owners without pet care challenges reported this possibility. Although these data must be interpreted with caution given the smaller number of ED patients surveyed, the stark contrast - combined with the higher likelihood of recurring pet care challenges for those with prior challenges during hospitalization - further spotlights value for early assessment of pet care responsibilities during the intake stages of hospitalization, and recognition of pet care challenges as a likely barrier to optimal medical outcomes. This point is supported by participant comments that collectively demonstrate the high value of animal companions who factor into delayed, deferred, or truncated care.

This work demonstrates a new facet to the importance of assessment of social support to determine who is most vulnerable to healthcare engagement barriers, similar to what has been documented in previous studies^19,20^. Inverse relationships between social support and pet care challenges in our study were noteworthy. Groups with lower social support included neurology and non-binary gender patients. Each group represents a potential “at-risk” cohort, who, along with patients who have emergent diagnoses or procedures, or previous pet care challenges, may be more likely to defer medical services or have compromised care^21^. This likely reflects a vulnerability due to social marginalization and health factors that may impact access and utilization of healthcare more broadly^22,23^. Patients who exhibit more than one of these risk factors may be particularly vulnerable, but interactions between these risk factors and hospital outcomes require further study.

Study strengths include objective EMR data and a prospective design to minimize recall bias. A nuanced evaluation of social support illuminated complex ways in which family, friends, and community networks contribute to patients’ ability to navigate pet and medical care. That said, some limitations should be noted. Pandemic restrictions slowed recruitment for approximately 2 years, potentially reducing power to detect rarer outcomes among subgroups.

The strength of bonding between participants and their pets was not assessed. Our sample was majority high-income and White, which may reduce generalizability. A higher frequency of multigenerational households and work-from-home situations post-pandemic could have created conservative estimates that may not reflect pre-pandemic times or future years as people return in-person work. Similarly, a high proportion of patients who endorsed availability of family, friends, or neighbors to assist with pet care may also not reflect the needs of other communities and may also have contributed to more conservative estimates regarding the scope of pet care needs and their impact.

In conclusion, this work provides new data regarding the role of animal companions in hospital-based courses and treatment among pet owners. Pet welfare appears to influence healthcare decisions and may play a key, but modifiable role in the care and disposition of ED and hospitalized patients, particularly those who lack sufficient social support. Future work that involves larger, representative samples is necessary to inform community-based programs to address this need.

## Acknowledgements

The authors would like to thank Michigan Humane for their assistance in this study, and Maddie’s Fund who provided gift funding to support these analyses.

## Data Availability Declaration

Data is provided within the manuscript or supplementary information files.

